# Early epidemiological assessment of the transmission potential and virulence of coronavirus disease 2019 (COVID-19) in Wuhan City: China, January-February, 2020

**DOI:** 10.1101/2020.02.12.20022434

**Authors:** Kenji Mizumoto, Katsushi Kagaya, Gerardo Chowell

## Abstract

**Background:** Since the first cluster of cases was identified in Wuhan City, China, in December, 2019, coronavirus disease 2019 (COVID-19) rapidly spread around the world. Despite the scarcity of publicly available data, scientists around the world have made strides in estimating the magnitude of the epidemic, the basic reproduction number, and transmission patterns. Accumulating evidence suggests that a substantial fraction of the infected individuals with the novel coronavirus show little if any symptoms, which highlights the need to reassess the transmission potential of this emerging disease. In this study, we derive estimates of the transmissibility and virulence of COVID-19 in Wuhan City, China, by reconstructing the underlying transmission dynamics using multiple data sources.

**Methods:** We employ statistical methods and publicly available epidemiological datasets to jointly derive estimates of transmissibility and severity associated with the novel coronavirus. For this purpose, the daily series of laboratory–confirmed COVID-19 cases and deaths in Wuhan City together with epidemiological data of Japanese repatriated from Wuhan City on board government–chartered flights were integrated into our analysis.

**Results:** Our posterior estimates of basic reproduction number (*R*) in Wuhan City, China in 2019–2020 reached values at 3.49 (95%CrI: 3.39–3.62) with a mean serial interval of 6.0 days, and the enhanced public health intervention after January 23^rd^ in 2020 was associated with a significantly reduced *R* at 0.84 (95%CrI: 0.81–0.88), with the total number of infections (i.e. cumulative infections) estimated at 1906634 (95%CrI: 1373500–2651124) in Wuhan City, elevating the overall proportion of infected individuals to 19.1% (95%CrI: 13.5–26.6%). We also estimated the most recent crude infection fatality ratio (IFR) and time–delay adjusted IFR at 0.04% (95% CrI: 0.03%–0.06%) and 0.12% (95%CrI: 0.08–0.17%), respectively, estimates that are several orders of magnitude smaller than the crude CFR estimated at 4.06%

**Conclusions:** We have estimated key epidemiological parameters of the transmissibility and virulence of COVID-19 in Wuhan, China during January-February, 2020 using an ecological modelling approach. The power of this approach lies in the ability to infer epidemiological parameters with quantified uncertainty from partial observations collected by surveillance systems.

## Background

The novel coronavirus (Severe acute respiratory syndrome coronavirus 2; SARS-CoV-2) that erupted from China is a deadly respiratory pathogen that belongs to the same family as the coronavirus responsible for the 2002-2003 Severe Acute Respiratory Syndrome (SARS) outbreaks [1]. Since the first cluster of cases was identified in Wuhan City, China, in December, 2019, the novel coronavirus disease 2019 (COVID-19) continues its relentless march around the world as of May 12^nd^, 2020 [2]. Nevertheless, China was hit hard by this emerging infectious disease, especially the city of Wuhan in Hubei Province, where the first cluster of severe pneumonia caused by the novel virus was identified. Meanwhile, the cumulative number of laboratory and clinically confirmed cases and deaths in mainland China has reached 82918and 4633, respectively, as of May 10^th^, 2020 [3].

Because the morbidity and mortality burden associated with the novel coronavirus has disproportionally affected the city of Wuhan, the center of the epidemic in China, the central government of the People’s Republic of China imposed a lockdown and social distancing measures in this city and surrounding areas starting on January 23^rd^ 2020. Indeed, out of the 82918 COVID-19 cases reported in China, 50339 cases (60.7%) are from Wuhan City. In terms of the death count, a total of 3869 deaths (83.5%) have been recorded in Wuhan city out of the 4633 deaths reported throughout China. To guide the effectiveness of interventions, it is crucial to gauge the uncertainty relating to key epidemiological parameters characterizing the transmissibility and the severity of the disease. Despite the scarcity of publicly available data, scientists around the world have made strides in estimating the magnitude of the epidemic, the basic reproduction number, and transmission patterns [4-5]. Moreover, accumulating evidence suggests that a substantial fraction of the infected individuals with the novel coronavirus show little if any symptoms, which suggest the need to reassess the transmission potential of this emerging disease [6]. For this purpose, in this study we employ statistical methods and publicly available epidemiological datasets to jointly derive estimates of transmissibility and severity associated with the novel coronavirus.

## Methods

### Epidemiological data

We linked our model to two different datasets. First, the daily series of laboratory–confirmed COVID-19 cases and deaths in Wuhan City were extracted according to date of symptoms onset or reporting date from several sources [3, 7-8]. Our analysis relies on epidemiological data reported prior to February 11^th^, 2020 because of the change in case definition that was announced on February 12th, 2020 [9]. As of February 11^th^, 2020, a total of 19559 confirmed cases including 820 deaths were reported in Wuhan City. Second, epidemiological data of Japanese evacuees from Wuhan City on board government–chartered flights were obtained from the Japanese government. After arriving in Japan, all of the Japanese evacuees were kept in isolation for about 14 days and examined for infection using polymerase chain reaction (PCR) tests [7]. As of February 11^th^, a total of four flights with the Japanese evacuees left Wuhan City. We collected information on the timing of the evacuee fights that left Wuhan City as well as the number of passengers that tested positive for COVID-19 in order to calibrate our model (Table S1).

### Statistical analysis

Using the following integral equation model, we estimate the reproduction number of COVID-19. Here, infected and reported cases are denoted by *i* and *c*, respectively.

We connected the daily incidence series with a discrete–time integral equation to describe the epidemic dynamics. Let *g_s_* denote the probability mass function of the serial interval, e.g., the time from illness onset in a primary case to illness onset in the secondary case, of length *s* days, which is given by

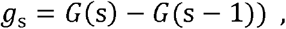

For *s* >0 where *G(.)* represents the cumulative distribution function of the gamma distribution. Mathematically, we describe the expected number of new cases with day *t*, E[*c*(t)] as follows,

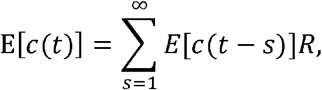

where E[*c*(t)] represents the expected number of new cases with onset day *t*, where *R* represents the average number of secondary cases per case.

Subsequently, we also employed the time–dependent variation in *R* to estimate the impact of enhanced interventions on the reproduction number. This time dependence was modelled by introducing a parameter *δ_1_*, which is given by

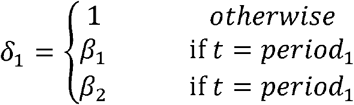

where period_1_ and period2 represent the corresponding period from January 23^rd^ to February 2^nd^ 2020 and from February 3^rd^ to February 11^th^, 2020, respectively. January 23^rd^ 2020 is the date when the central government of the People’s Republic of China imposed a lockdown in Wuhan and other cities in Hubei in an effort to quarantine the epicentre of the coronavirus (COVID-19) to mitigate transmission. Furthermore, we evenly divide the interval into two periods to incorporate the time-dependent effects on *R* using the parameters *β*_1_ and *β*_2_ which scale the effects of the intervention, taking values smaller than 1[10].

To account for the probability of occurrence, *θ* [11], we assume that the number of observed cases on day *t*, *h(t)*, occurred according to a Bernoulli sampling process, with the expected values E(*c_t_*;*H*_t−1_), where E(*c_t_*; *H*_t−1_) denotes the conditional expected incidence on day *t*, given the history of observed data from day 1 to day (*t*−1), denoted by *H*_t−1_. Thus, the number of expected newly observed cases is written as follows:

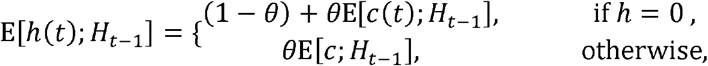

Further, we model the time–dependent variation in the reporting probability. This time dependence was modelled by introducing a parameter *δ_2_*, which is given by

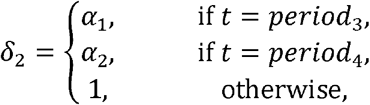

where period_3_ and period_4_ represent the corresponding periods from the start of our study period to Jan 16 and from Jan 17 to Jan 22, respectively, while *α_1_* and *α_2_* scale the extent of the reporting probability (where *α_1_* and *α_2_* is expected to be smaller than 1). We evenly divide the time interval before the lockdown was put in place into two periods in order to incorporate the time dependency of the reporting probability. The number of expected newly observed cases should be updated as

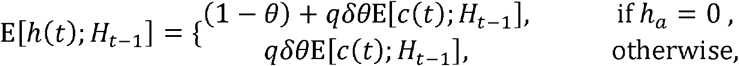

We assume the incidence, *h*(t) is the result of the Binomial sampling process with the expectation E[*h*]. The likelihood function for the time series of observed cases that we employ to estimate the effective reproduction number and other relevant parameters is given by:

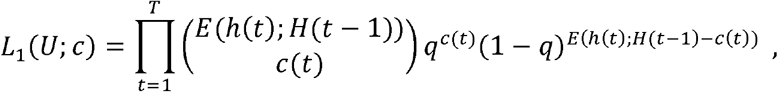

where *U* indicates parameter sets that are estimated from this likelihood.

Subsequently, the conditional probability of non–infection given residents in Wuhan City at the time point of *t_i_*, *p_ti_*, was assumed to follow a binomial distribution, and the likelihood function is given by:

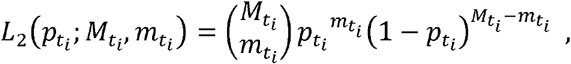

Where *M*_ti_ and *m*_ti_ is the number of government charted flight passengers and non–infected passengers at the date of *t*_i_, respecitively, and *p*_ti_ is the proportion of the estimated non–infected population in Wuhan at the date of *t*_i_, calculated from the *h*(t) and catchment population in Wuhan City [3,13].

Serial interval estimates of COVID-19 were derived from previous studies of COVID-19, indicating that it follows a gamma distribution with the mean and SD at 6.0 and 2.9 days, respectively, based on ref. [14,15]. The maximum value of the serial interval was fixed at 28 days as the cumulative probability distribution of the gamma distribution up to 28 days reaches 1.000.

### Infection fatality ratio

Crude CFR and crude IFR is defined as the number of cumulative deaths divided by the number of cumulative cases or infections at a specific point in time without adjusting the time delay from illness onset or hospitalization to death. Next, we employed an integral equation model in order to estimate the real–time IFR. First, we estimated the real–time CFR as described elsewhere [16-18]. For the estimation, we employ the delay from hospitalization to death, *f*_s_, which is assumed to be given by *f*_s_ = *F*(s) − *F*(s−1) for *s*>0 where *H*(s) follows a gamma distribution with mean 10.1 days and SD 5.4 days, obtained from the available observed data [19].

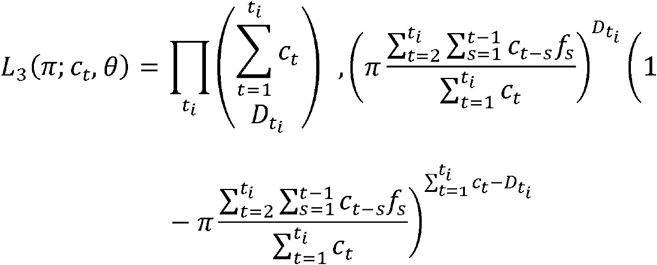

where *c_t_* represents the number of new cases with reported day *t*, and *D_ti_* is the number of new deaths with reported day *t*_i_ [16-18]. We assume that the cumulative number of observed deaths, *D*_t_ is the result of the binomial sampling process with probability *π*. Subsequently, crude IFR and time–delay adjusted IFR are calculated using the estimated *π* and *h*_t_.

The total likelihood is calculated as *L*=*L*_1_*L*_2_*L*_3_ and model parameters were estimated using a Monte Carlo Markov Chain (MCMC) method in a Bayesian framework. Posterior distributions of the model parameters were estimated based on sampling from the three Markov chains. For each chain, we drew 100,000 samples from the posterior distribution after a burn–in of 20,000 iterations. Convergence of MCMC chains were evaluated using the potential scale reduction statistic [20-21]. Estimates and 95% credibility intervals for these estimates are based on the posterior probability distribution of each parameter and based on the samples drawn from the posterior distributions. All statistical analyses were conducted in R version 3.5.2 (R Foundation for Statistical Computing, Vienna, Austria) using the ‘rstan’ package.

## Results

The daily series of COVID-19 laboratory–confirmed incidence and cumulative incidence in Wuhan in 2019–2020 are displayed in Figure 1. Overall, our dynamical models yield a good fit to the temporal dynamics (i.e. incidence, cumulative incidence) including an early exponential growth pattern in Wuhan. In incidence data, a few fluctuations are evident, probably indicating that the surveillance system likely missed many cases during the early transmission phase (Figure 1).

**Figure 1.**
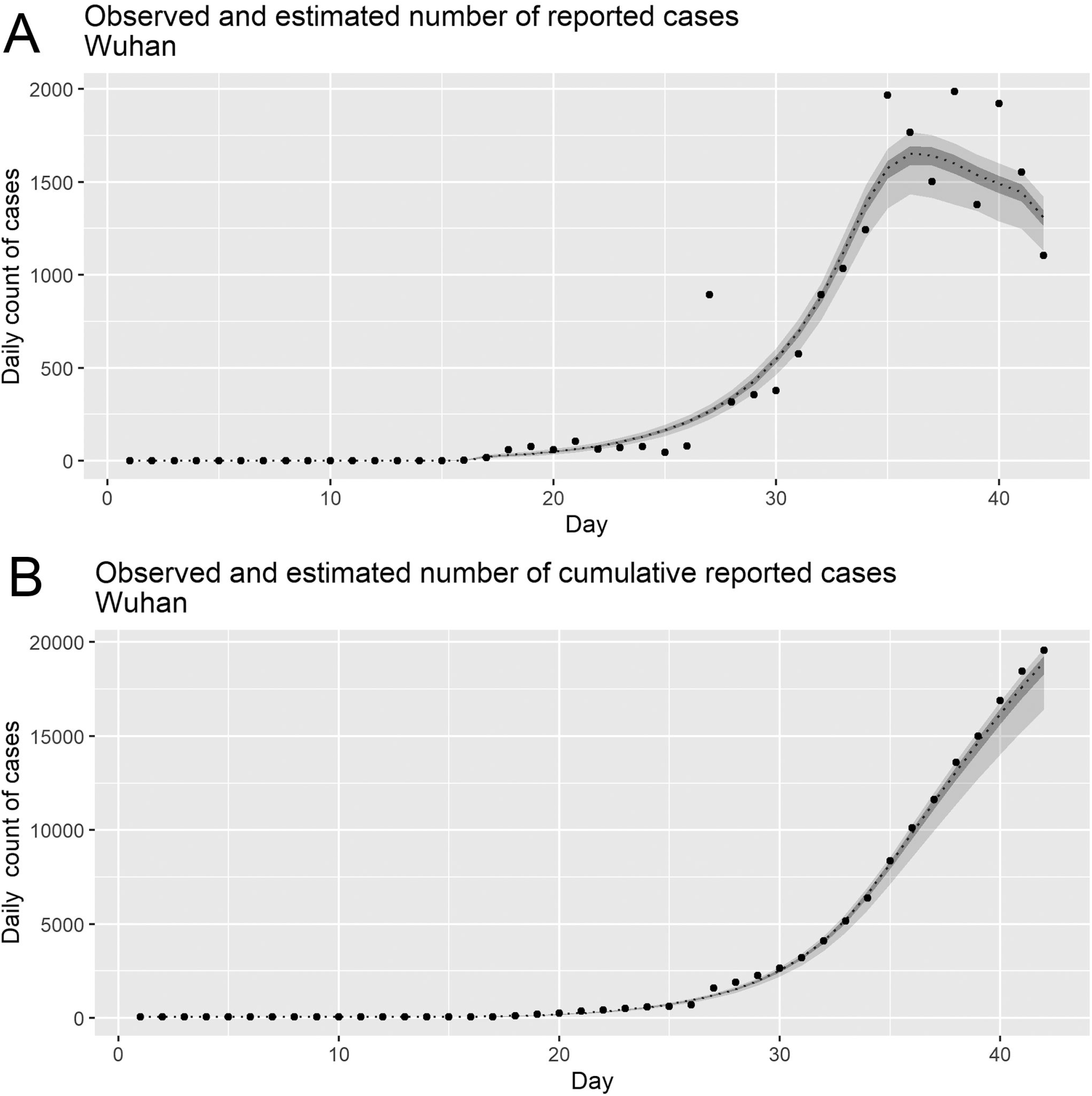
Observed and posterior estimates of the daily new cases and cumulative cases of the COVID-19 cases in Wuhan, China, 2019–2020. Observed and posterior estimates of laboratory–confirmed reported cases (A) and cumulative reported cases (B) are presented. Observed data are presented in the dot, while dashed line indicates 50 percentile, and areas surrounded by light grey and deep grey indicates 95% and 50% credible intervals (CrI) for posterior estimates, respectively. Epidemic day 1 corresponds to the day that starts at January 1^st^, 2020.

Our posterior estimates of basic reproduction number (*R*) in Wuhan City, China in 2019–2020 was estimated to be 3.49 (95%CrI: 3.39–3.62). The time–dependent scaling factors quantifying the extent of enhanced public health intervention on *R* is 0.99 (95%CrI: 0.95–1.00), declining *R* to 3.44 (95%CrI: 3.36–3.52) from January 23^rd^ to February 1^st^ and 0.24 (95%CrI: 0.23–0.26), declining *R* to 0.84 (95%CrI: 0.81–0.88) from February 2^nd^ to February 11^th^, 2020. Other parameter estimates for the probability of occurrence and reporting rate are 0.97 (95% CrI: 0.84–1.00) and 0.010 (95% CrI: 0.007–0.014), respectively. Moreover, the time–dependent scaling factor quantifying the extent of reporting rate, *α*, is estimated to be 0.07 (95% CrI: 0.03–0.18) before January 16^th^ and to be 0.99 (95% CrI: 0.96–1.00) from January 17^th^ to January 22^nd^.

We conducted sensitivity analyses to examine how varying the mean serial interval between 5.0 and 7.0 days affects our *R* estimates. *R* estimates are sensitive to changes in the serial interval, ranging from 2.86 (95%CrI: 2.79–2.96) to 4.10 (95%CrI: 3.96–4.38).

The total number of estimated laboratory–confirmed cases (i.e. cumulative cases) is 18967 (95% CrI: 16428–19680) while the actual numbers of reported laboratory–confirmed cases during our study period is 19559 as of February 11th, 2020. Moreover, we inferred the total number of COVID–19 infections (Figure S1). Our results indicate that the total number of infections (i.e. cumulative infections) is 1906634 (95%CrI: 1373500–2651124).

The Observed and posterior estimates of the cumulative number of deaths from COVID–19 in Wuhan are displayed in Figure 2, and model–based posterior estimates of the cumulative number of deaths is 821 (95%CrI: 751–892), while actual number of reported deaths is 820. The estimated temporal variation in the death risk caused by COVID–19 in Wuhan, China, 2019–2020 is shown in Figure 3 and Figure S2. Observed and posterior estimates of the crude CFR in Wuhan City is presented in Figure 2A, while observed and posterior estimates of time–delay adjusted CFR is shown in Figure 2B. Furthermore, Figure 3A and 3B illustrates time–delay no–adjusted IFR and time–delay adjusted IFR, respectively.

**Figure 2.**
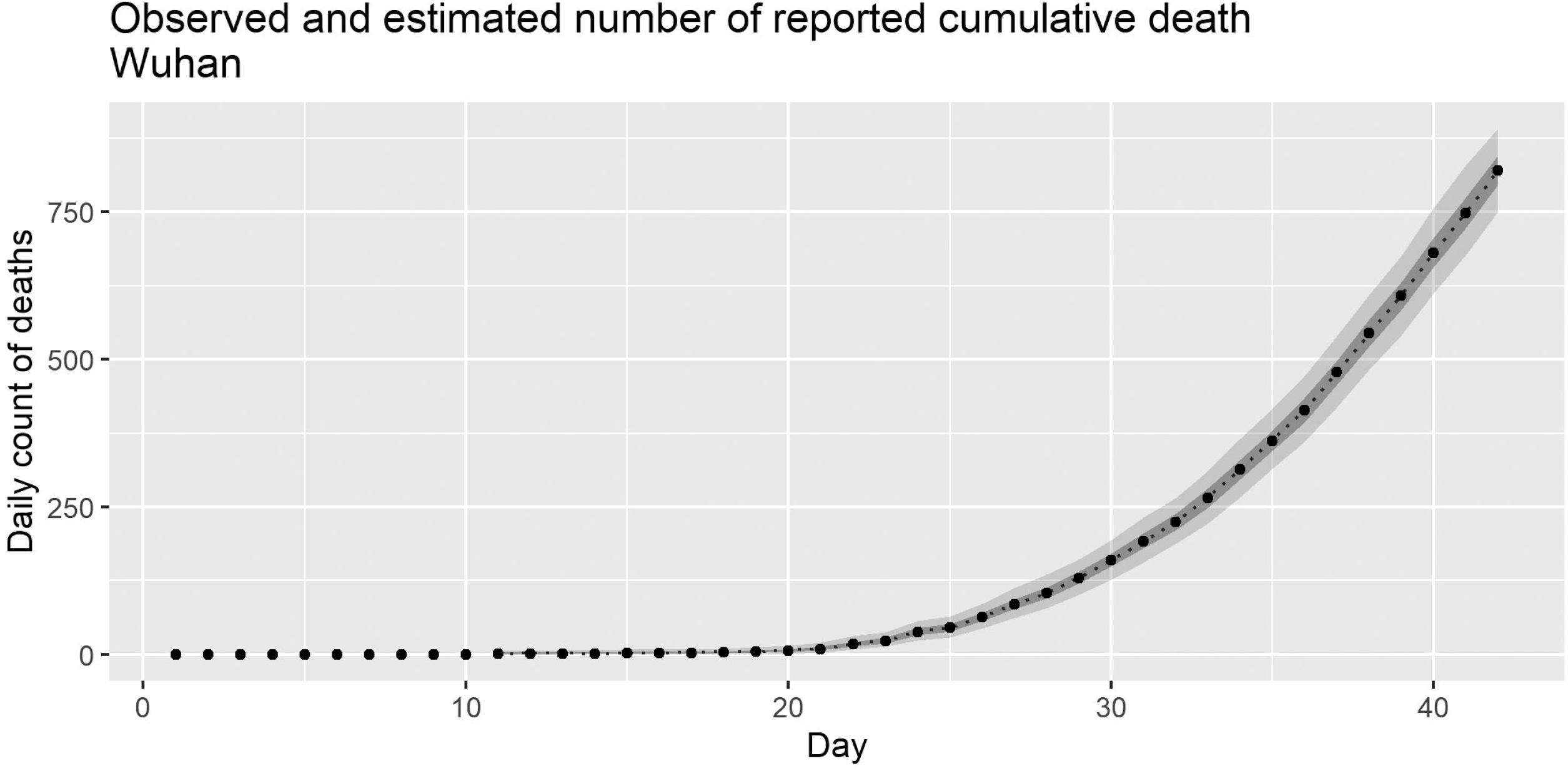
Observed and posterior estimates of the cumulative deaths of the COVID-19 in Wuhan, China, 2019–2020. Observed and posterior estimates of the cumulative deaths of the COVID-19 in Wuhan is presented. Observed data are presented in the dot, while dashed line indicates 50 percentile, and areas surrounded by light grey and deep grey indicates 95% and 50% credible intervals (CrI) for posterior estimates, respectively. Epidemic day 1 corresponds to the day that starts at January 1^st^, 2020.

**Figure 3.**
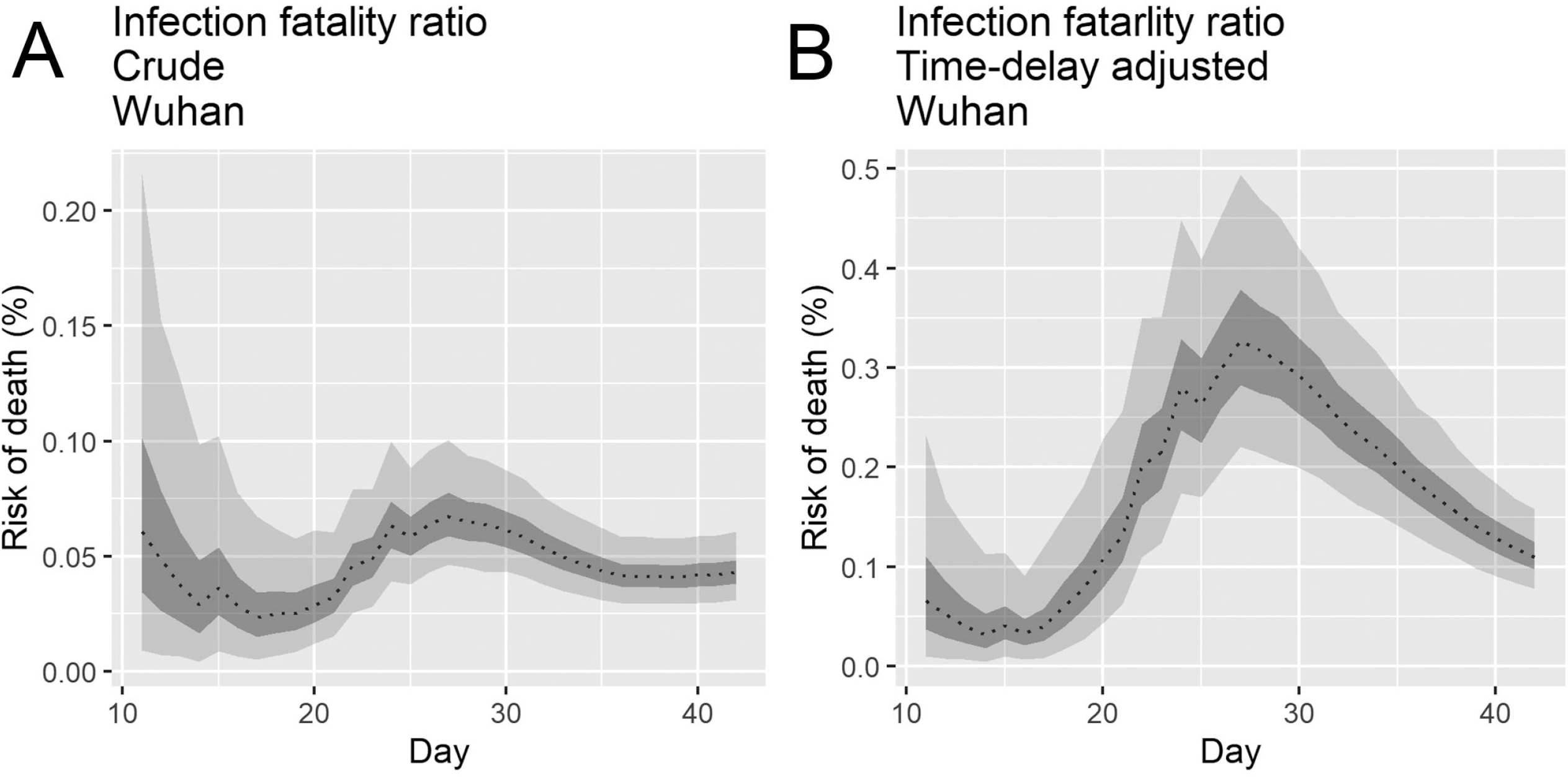
Temporal variation of the infection fatality risks caused by COVID-19 in Wuhan, China, 2019–2020. (A) Posterior estimates of crude infection fatality ratio in Wuhan City. (B) Posterior estimates of time–delay adjusted infection fatality ratio in Wuhan City. Black dots shows observed data, and light and dark indicates 95% and 50% credible intervals for posterior estimates, respectively. Epidemic day 1 corresponds to the day that starts at January 1^st^, 2020.

The latest estimate of the crude CFR and time–delay adjusted CFR in Wuhan appeared to be 4.3% (95% CrI: 3.9–5.0%) and 12.2% (95% CrI: 11.4–13.1%), respectively, whereas the latest model–based posterior estimates of time–delay not adjusted IFR and adjusted IFR, presented in Figure 3 C and D, are 0.04%(95% CrI: 0.03%-0.06%) and 0.12% (95%CrI: 0.08–0.17%), respectively, while the observed crude CFR is calculated to be 4.06% (Table 1).

**Table 1.** – Death risk by COVID-19 in Wuhan City, China, 2020 (As of February 12, 2020)

| Death Risk | Latest estimate | Range of median estimates |
| --- | --- | --- |
| Crude CFR (Observed) | 4.06% | 2.0 – 9.0% |
| Crude CFR (Estimated) | 4.3% (95%CrI<sup>‡</sup>: 3.9 – 5.0%) | 3.4 – 7.1% |
| Time delay adjusted CFR | 12.2% (95%CrI: 11.4 – 13.1%) | 4.0 – 34.5% |
| Crud IFR | 0.04% (95%CrI: 0.03 – 0.06%) | 0.02 – 0.07% |
| Time delay adjusted IFR | 0.12% (95%CrI: 0.08 – 0.17%) | 0.03 – 0.33% |

CrI: Credibility intervals, CFR: Case fatality ratio, IFR: Infection fatality ratio
<sup>‡</sup>Upper and lower 95% credibility interval

## Discussion

In this study we derived estimates of the transmissibility and virulence of COVID-19 in Wuhan City, China, by reconstructing the underlying transmission dynamics using multiple data sources. Applying dynamic modeling, the reproduction number, death risks as well as probabilities of occurrence and reporting rate were estimated.

Our posterior estimates of basic reproduction number (R) in Wuhan City, China in 2019–2020 is calculated to be 3.49 (95%CrI: 3.39–3.62). The time–dependent scaling factor quantifying the extent of enhanced public health intervention on *R* is 0.99 (95%CrI: 0.95–1.00), declining *R* to 3.44 (95%CrI: 3.36–3.52) from January 23^rd^ to February 1^st^ and a scaling factor at 0.24 (95%CrI: 0.23–0.26), declining *R* to 0.84 (95%CrI: 0.81–0.88) for February 2^nd^ to February 11^th^, 2020. These *R* estimates capturing the underlying transmission dynamics modify the impact of COVID-19, with the total number of infections (i.e. cumulative infections) estimated at 1906634 (95%CrI: 1373500-2651124) in Wuhan City, raising the proportion of infected individuals to 19.1% (95%CrI: 13.7–26.5%) with a catchment population in Wuhan City of 10 million people. Our estimates of mean reproduction number reached values of 3.44, an estimate consistent with previous mean estimates in the range 2.2–3.8 derived by fitting epidemic models to the initial growth phase of the observed case incidence [14,22,23]. By comparison, the *R* estimate for the Diamond Princess cruise ship in Japan reached values as high as ~11 [24]. Further, these estimates are higher than recent mean *R* estimates derived from the growth rates of the COVID-19 outbreaks in Singapore (R~1.1) [25] and Korea (R~1.5) [26].

The sustained high *R* values in Wuhan City even after the lockdown and mobility restrictions suggests that transmission continues inside the household or amplified in healthcare settings [19], which is a landmark of past SARS and MERS outbreaks [27-28]. Considering the potent transmissibility of COVID-19 in confined settings, as illustrated by COVID-19 outbreaks aboard cruise ships, including the Diamond Princess cruise ship, where the total number of secondary or tertiary infections reached 705 among more than 3,700 passengers as of February 28^th^, 2020 and also by the COVID-19 outbreak tied to the Shincheonji religious sect in South Korea where church members appear to have infected from seven to 10 people [29-31], it is crucial to prevent transmission in confined settings including hospital-based transmission by strengthening infection control measures as well as transmission stemming from large social gatherings.

Our most recent estimates of the crude CFR and time–delay adjusted CFR for Wuhan city are at 4.3% (95% CrI: 3.9–5.0%) and 12.2% (95% CrI: 11.4–13.1%), respectively. In contrast, our most recent crude IFR and time–delay adjusted IFR is estimated to be 0.04% (95% CrI: 0.03%-0.06%) and 0.12% (95%CrI: 0.08–0.17%), which is several orders of magnitude smaller than the crude CFR estimated at 4.06% and another recent estimate of the infection fatality ratio at 0.66% (95%CrI: 0.39–1.33) and 0.6% (95% CI: 0.2–1.3) in China [32, 33]. Several data and methodological differences can explain these differences, which we list in Table S2. For instance, Verity et al. conducts an age adjustment based on the data of age-stratified COVID-19 deaths from mainland China, assumes a constant attack rate by age and adjusts for demographic structure. Our IFR estimates will be compared with estimates emerging from ongoing several mass serological studies in China (Wuhan City), Italy, Germany the U.K., and New York. Yet, these serological studies should be carefully validated since these are not exempt of limitations as discussed elsewhere [34, 35]. Also, these findings indicate that the death risk in Wuhan is estimated to be much higher than those in other areas, which is likely explained by hospital-based transmission [36]. Indeed, past nosocomial outbreaks have been reported to elevate the CFR associated with MERS and SARS outbreaks, where inpatients that tend to be older and affected by underlying diseases have raised the CFR to values as high as 20% for a MERS outbreak [37-38].

Public health authorities are interested in quantifying both *R* and CFR to measure the transmission potential and virulence of an infectious disease, especially when emerging/re–emerging epidemics occur in order to decide the intensity of the public health response. In the context of a substantial fraction of unobserved infections due to COVID-19, *R* estimates derived from the trajectory of infections and the IFR are more realistic indicators compared to estimates derived from observed cases alone [18, 39-40].

Our analysis also revealed a high probability of occurrence and quite low reporting probabilities in Wuhan City. High probability of occurrence in the above equation suggests that zero observed cases at some point is not due to the absence of those infected, but more likely due to a low reporting rate. A very low reporting probability suggests that it is difficult to diagnose COVID-19 cases or a breakdown in medical care delivery. Moreover, we also identified a remarkable change in the reporting rate, estimated to be 14–fold lower in the 1^st^ period (–Jan 16^th^, 2020) and about the same during the 2^nd^ period (January 17^th^ – 22^nd^, 2020), relative to that estimated after January 23^rd^ 2020.

Our results are not free from the limitations. First, our methodology aims to capture the underlying transmission dynamics using multiple data sources. By implementing mass screening in certain populations is a useful approach to ascertain the real proportion of those infected and a way of adding credibility to the estimated values. Second, it is worth noting that the data of Japanese evacuees from Wuhan employed in our analysis is not a random sample from the Wuhan catchment population. Indeed, it also plausible that their risk of infection in this sample is not as high as that of local residents in Wuhan, underestimating the fatality risk. Third, given the likely under-ascertainment of cases, there may also exist unreported deaths, and this might underestimate the death risk. Fourth, case fatality ratio (CFR) varies with age, gender, presence or absence of comorbidities, race, whether the healthcare system is overloaded or not, and other factors such as poverty risk, infant mortality risk, and the cumulative morbidity ratio [41-45]. As CFR is influence by reporting rate and ascertainment bias, subgroup analysis of IFR based on individual-level data is essential to capture the overall disease burden of COVID-19.

## Conclusion

In summary, we have estimated key epidemiological parameters of the transmissibility and virulence of COVID-19 in Wuhan, China, January-February, 2020 using an ecological modelling approach and several epidemiological datasets. The power of our approach lies in the ability to infer epidemiological parameters with quantified uncertainty from partial observations collected by surveillance systems.

## Data Availability

The present study relies on published data and access information to essential components of the data are available from the corresponding author.

## List of abbreviations

CFR: Case fatality ratio,
IFR: Infection Fatality ratio,
SARS: Severe Acute Respiratory Syndrome,
MERS: Middle East Respiratory Syndrome

### Additional files

**Additional file 1: Table S1**. Information related to Japanese evacuees from Wuhan City on board government–chartered flights. **Table S2**. Main differences between our study and previous study.

**Additional file 2: Fig. S1. Observed daily new cases and posterior estimates of the daily new infections of the COVID-19 in Wuhan, China, 2019–2020**.

Observed daily new cases and posterior estimates of infections of the COVID-19 are presented. Observed data are presented in the dot, while dashed line indicates 50 percentile, and areas surrounded by light grey and deep grey indicates 95% and 50% credible intervals (CrI) for posterior estimates, respectively. Epidemic day 1 corresponds to the day that starts at January 1^st^, 2020.

**Additional file 3: Fig. S2. Temporal variation of the case fatality risks caused by COVID-19 in Wuhan, China, 2019–2020**.

(A) Observed and posterior estimates of crude case fatality ratio in Wuhan City, (B) Observed crude case fatality ratio and posterior estimates of time–delay adjusted CFR in Wuhan City. This figure is submitted to the ref [19]. The purpose of the study is to compare the case fatality ration (CFR. Not IFR) in three different areas (Wuhan City, in Hubei Province excluding Wuhan City and in China excluding Hubei Province) to interpret the current severity of the epidemic in China, and the purpose is different from this study.

## Declarations

### Ethics approval and consent to participate

Not applicable.

### Consent for publication

Not applicable.

### Authors’ contributions

KM and GC conceived the early study idea. KM and KK built the model. KM implemented statistical analysis and wrote the first full draft. GC advised on and helped shape the research. All authors contributed to the interpretation of the results and edited and commented on several earlier versions of the manuscript. All authors read and approved the final manuscript.

## Acknowledgements

Not applicable.

